# A Computational Model for Estimating the Evolution of COVID-19 in Rondônia-Brazil

**DOI:** 10.1101/2020.05.05.20091942

**Authors:** Tito Dias Junior, Camila Bueno Machado

## Abstract

In this work, the modified SEIR model was proposed to account separately for the tested and isolated cases, with severe and critical symptoms, from those not tested, with mild and moderate symptoms. Two parameters were estimated and evaluated for the cases registered in Rondônia, Brazil, between March 20 and April 22. The basic reproduction rate did not remain constant during the period, showing eventual variations due to social behavior. The results show that an increase in the proportion of testing to about 56% provided a significant decrease in confirmed cases, for the expansion of tested cases beyond the current testing criterion (20%) would help to identify and isolate also mild and moderate cases, generally referred to as asymptomatic.

## 1. Introduction

In December 2019, COVID-19 (Coronavirus disease 2019), caused by SARS-CoV-2, emerged in Wuhan, China, and, despite the extensive and severe containment measures implemented by the Chinese government, it disseminated to other regions, having currently reached several countries on all continents [1]. In Brazil, the first confirmed case for COVID-19 was a 61-year-old male patient with a history of travel from Italy registered on 02/25/2020 in São Paulo-SP.

In the Brazilian state of Rondônia, the first confirmed case was registered on 03/20/2020: a patient with history of travel from São Paulo/SP to Ji-Paraná/RO [2]. On the same date, the State Government of Rondônia published the Decree No. 24,887 of March 20 2020, that implemented comprehensive measures of social isolation [3]. In this work, we have utilized the official data made available by ANVISA (National Health Surveillance Agency) regarding Rondônia from 03/20/2020 to 04/22/2020 in the study of the proposed mathematical model to estimate epidemiological parameters, which are related to behavior population in response to social isolation measures enacted by the State Government. The present study aimed to propose a mathematical model that represents the quantitative epidemiological evolution of COVID-19 in Rondônia, as well as to evaluate the influence of social behavior parameters and testing criteria on the number of confirmed cases and, consequently, on the need for hospitalizations of severe and critical cases and the mortality rate.

## 2. Methods

### 2.1. Mathematical Modeling

For the modeling carried out in this work, we have proposed a modification of the SEIR model (Susceptible-Exposed-Infected-Removed) [4–6]. In such modified model, the number of susceptible individuals S decreases according to the rate of effective contacts between the untested infected and the susceptible (β), assuming that the tested infected remain isolated according to the guidelines of the health authorities [7–9]. The number of individuals exposed *E* increases in proportion to the decrease in susceptible individuals and decreases with the proportion that those exposed become infected after the average incubation period (1/*α*). In this model, unlike traditional SEIR modeling, we chose to classify the infected, recovered and deceased in two groups, namely, tested and untested groups. Thus, the parameter *ρ* is introduced to quantify the proportion of individuals tested, so that in the computer simulation phase, it may be possible to evaluate the effect of the testing strategy on the results obtained. Therefore, the number of infected individuals tested will increase with the number of exposed individuals who have undergone incubation and decrease with those who have recovered or died, according to the mortality rate *c* and the average period of infection 1/*γ*. In the obtained equation, it can be demonstrated, using the method of the subsequent generation matrix [4], that the basic rate of reproduction is defined by the *R*_0_ (*t*) = *β/γ*; it may vary over time since it is assumed that the rate of effective contacts (*β*) depends on the strategy of social isolation or the use of collective masks. The total effective population considered in modeling viral transmission can be expressed by *N = S + E + I_t_ + I_nt_ + R_t_ + R_nt_*, where *S* corresponds to susceptible individuals, *E* to individuals exposed to the virus, *I_t_* and *I_nt_* to the tested and untested infected, respectively, and *R_t_* and *R_nt_* to the tested and untested recoveries; furthermore, those who do not recover die (*D_t_* and *D_nt_*). Thus, the mathematical model can be formulated as:

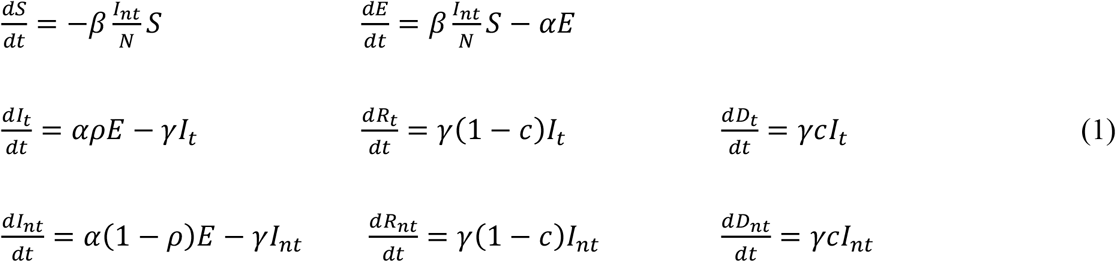

### 2.2. Numerical Solution

For the numerical solution of the system of eight ordinary differential equations resulting from the mathematical modeling of the problem, we employed the 4th order Runge-Kutta algorithm. For this, the Python programming language was used, through the Numpy libraries [10] and *Scipy* [11]. The algorithm for the integration step keeps the local error of integration limited by a linear combination of the relative (10^−3^) and absolute tolerances (10^−6^). All initial values for the variables are null, except for the number of susceptible individuals that is equal to the population in 2019 estimated by the IBGE (Brazilian Institute of Geography and Statistics) for the region under study minus the number of untested cases that was one. Thus, we considered a healthy population fully susceptible to have had initial contact with only one infected and untested case. We obtained the system solutions for some sets of parameters corresponding to each analyzed case, considering, for example, the initial population of susceptible individuals, the mortality rate, the proportion of individuals tested, and other parameters, according to the hypothetical case established.

### 2.3. Case Study

To estimate the evolution of COVID-19 cases in Rondônia, the official data published by ANVISA (National Health Surveillance Agency) on 04/22/2020 [12], containing the numbers of new and accumulated cases and new and accumulated deaths, were used. The initial number of susceptible individuals was considered equal to the population of Rondônia estimated by the IBGE for 2019 [13]. We estimate the proportion of tested and untested cases at *ρ* = 0.2 and the mortality rate at *c* = 0.02, according to official data for Rondônia. To simulate the statistical dispersion of the average periods of incubation (1*/α*) and infection (1/*γ*), the mean values suggested in the literature, that is, 1/*γ* = 3 and 1/*α* = 5.1, were used in the Erlang distribution with shape equal to two, obtaining periods greater than or equal to one variable around the suggested means [6,14,15]. The model estimated the rate of effective contacts from the basic reproduction rate and the period of infection (*β* = *R*_0_*γ*).

We performed the simulations for three values of the basic reproduction rate *R*_0_ admitted constant over time and another with variable *R*_0_ with *t*, as a result of changes in social isolation behavior or the collective use of masks, allowing intervals for *R*_0_(*t*) or phases. For variable *R*_0_(*t*), we evaluated the influence of the proportion of testing on the number of accumulated known cases (*I_t_* + *R_t_ + D_t_ + D_nt_*), that is, the influence of the relationship between the number of infected and tested individuals. It is noteworthy that in this modeling, the death count of infected individuals not tested should be added to the accumulated cases since, in practice, after death under suspicion, these individuals undergo testing confirming the infection, making them known. For each set of parameters, we have performed the simulations 100 times.

## 3. Results e Discussion

As in all mathematical modeling, values should be estimated or arbitrated for model parameters based on simplifying hypotheses and scientific evidence. In addition to those parameters, defined in Section 2.3, obtained from estimates in the literature, the proportion of individuals tested was, initially, estimated at 20% (*ρ* = 0.2), that is, the clinical and epidemiological testing criteria regularly include 20% of all infected individuals in the studied region. In Rondônia, according to the daily bulletin of the State Health Agency [16], as of 04/21/2020, there were 223 infected, 1220 discarded tests and 115 awaiting results, totaling 1558 tests performed, including 4 deaths. The criterion established in Rondônia requires testing for cases that present clinical symptoms: fever with at least one respiratory symptom and one of the epidemiological criteria – a history of travel to an area with local transmission or history of close contact with a suspected case of COVID-19, in the 14 days before the onset of symptoms [17]. In this context, the WHO (World Health Organization) estimated that, in China, 80% of confirmed patients developed mild to moderate symptoms, 13.8% had severe symptoms, and 6.1% critical symptoms [18]. Therefore, considering the testing criteria used and the incidence of severe and critical cases mentioned, we estimated the proportion of those tested at 20% (*ρ* = 0.2). The estimated lethality rate was 2%. To assess the influence of the basic reproduction rate on the adequacy of the proposed theoretical model to confirmed cases registered in Rondônia, we simulated the mathematical model for 40 days for three situations with different arbitrated values for the basic reproduction rate (*R*_0_) constant during the simulated period, the first day was that with the appearance of the first confirmed case.

In Figure 1, the higher the value of *R*_0_, the greater the number of cases accumulated during the simulated period. Official data for confirmed cases are shown [12], and the proposed model captured its exponential trend, demonstrating in general terms the adequacy of the model to the registered reality. However, the simulations performed for *R*_0_ constant in the considered time interval do not fit precisely with the data, suggesting that the basic reproduction rate did not remain constant in the recorded period. Figure 2 shows the same simulations and real data for the logarithmic scale of cases, displaying the slopes of the curves from the simulations and the variations in the slope of the recorded data. Note that until the 15th day the slope of the trend of confirmed cases tends to approach the curve *R*_0_ = 2.2, between the 15th and the 25th, it approaches the curve *R*_0_ = 2.3, after the 25th day it approaches the curve *R*_0_ = 2.4 until the 31st day, and then following the trend of approaching the curve *R*_0_ = 2.6.

**Figure 1:**
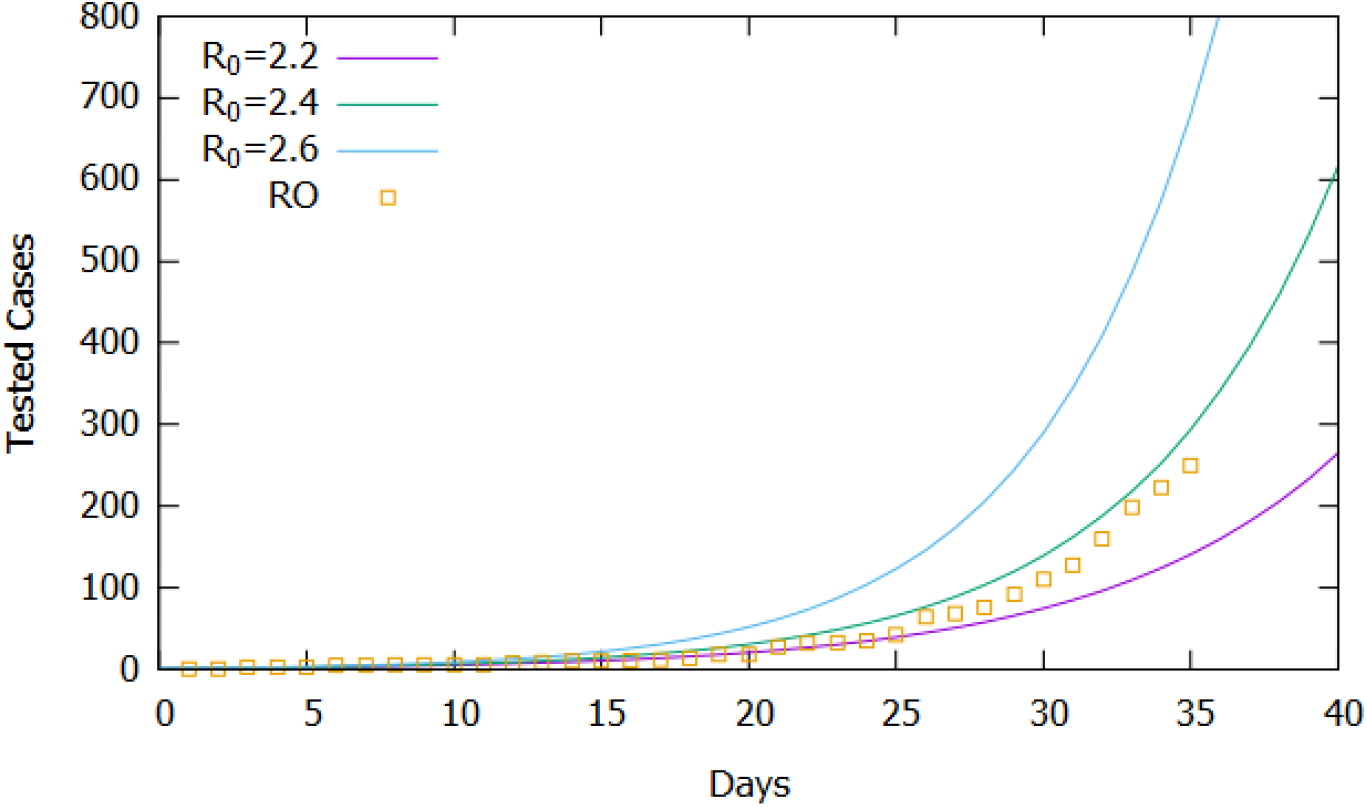
*Comparison of simulated results for constant values of R_0_ and confirmed cases in Rondônia (04/22/2020)*.

**Figure 2:**
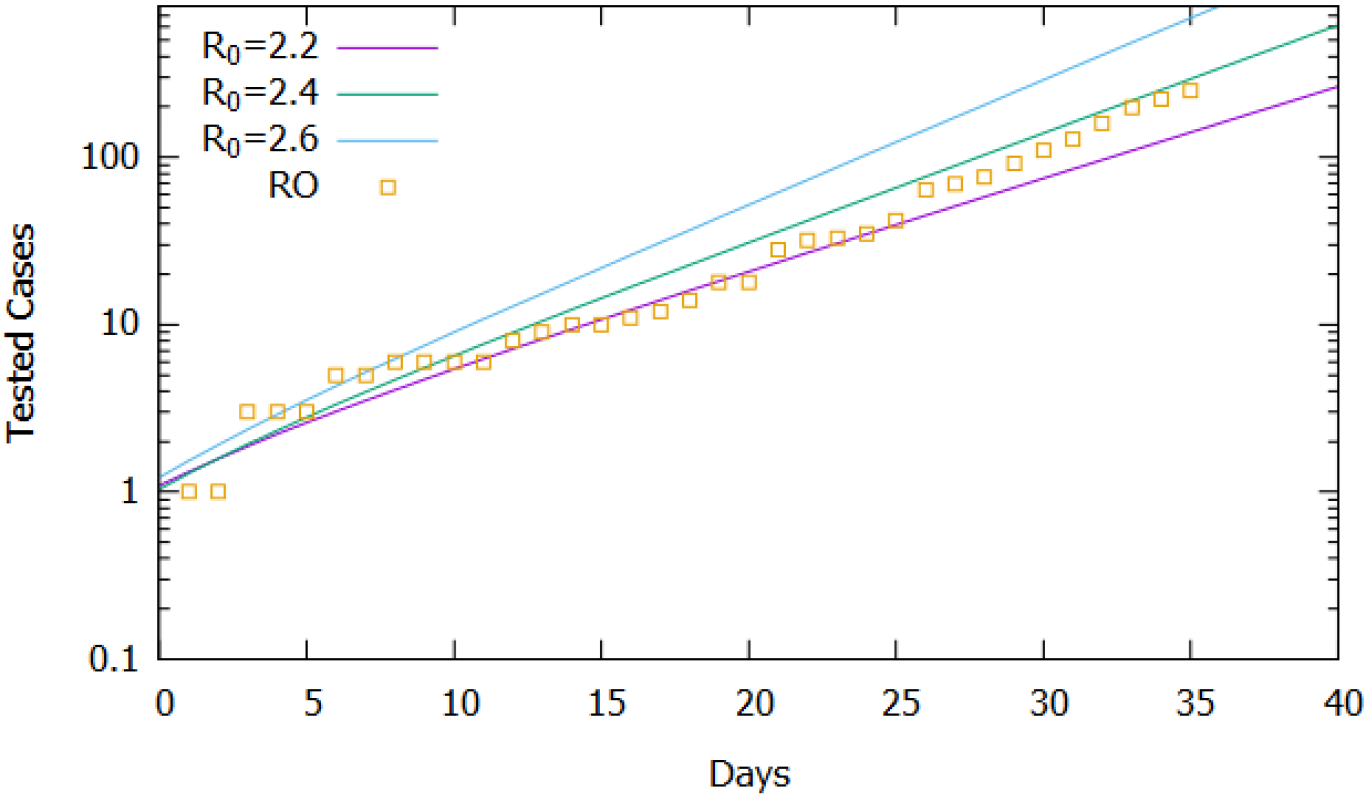
*Comparison of simulated results for constant values of R_0_ and confirmed cases in Rondônia (04/22/2020) on a logarithmic scale for the cases*.

From the analysis of Figure 2, we could roughly estimate the values for the basic reproduction rate as varying in the period considered according to eq. (2), which establishes four intervals or phases. Figure 3 shows, in a logarithmic scale, that the simulation result approximates more accurately to the recorded data, validating the estimate of the variation of the basic reproduction rate in the considered interval. It is important to note that the rate of effective contacts derives from the basic reproduction rate and the period of infection (*β* = *R*_0_*γ*), therefore, for a given period of infection, a change in the rate of effective contacts causes a directly proportional variation basic reproduction rate. In this context, changes in social behavior, whether through isolation or the use of homemade masks, can decrease the rate of effective contacts and consequently interfere with the basic reproduction rate of COVID-19.

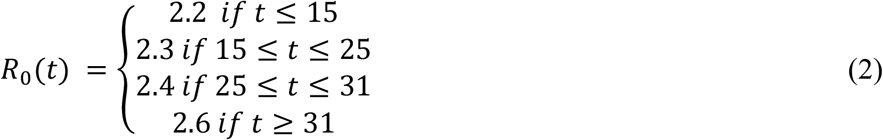

**Figure 3:**
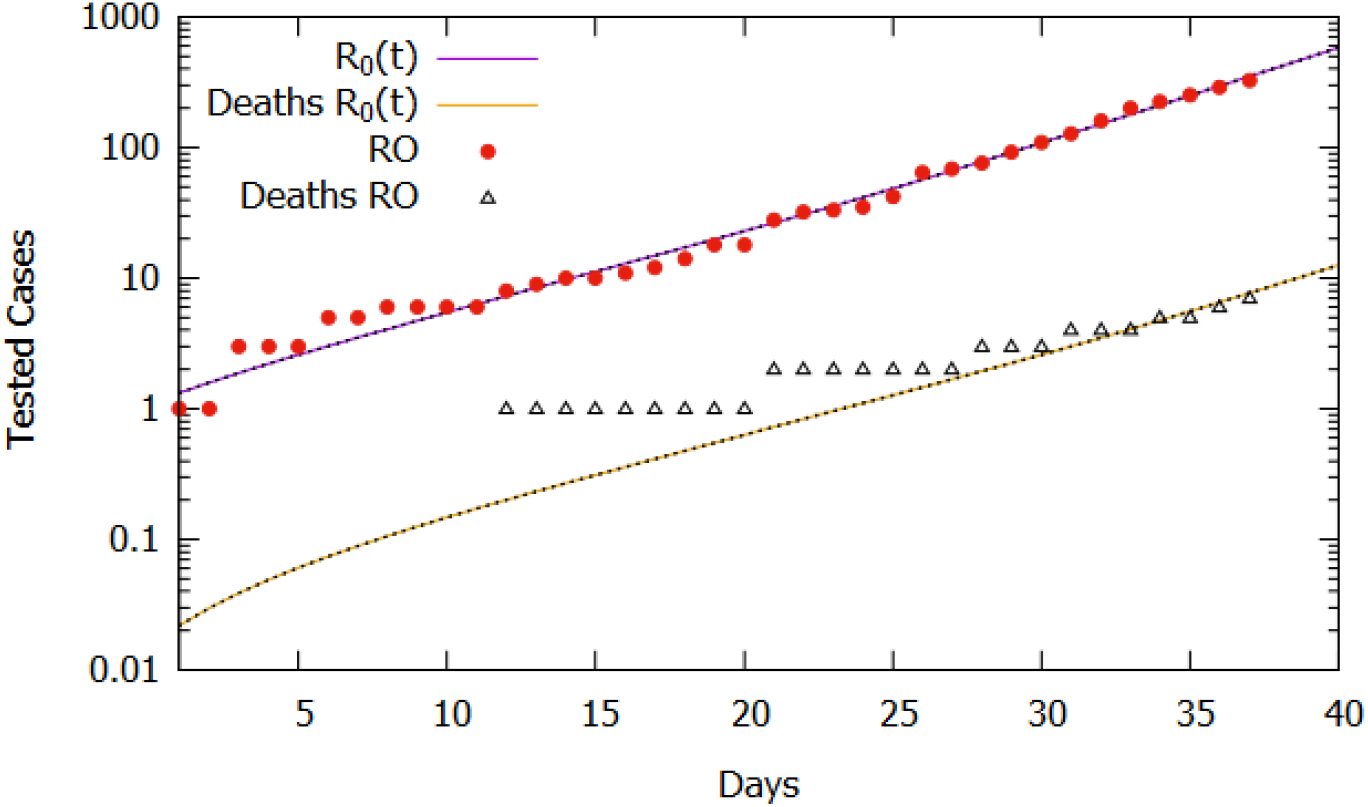
*Comparison between confirmed cases and deaths and the simulation for R_0_(t) according to eq. (2)*.

Considering the estimate for the function *R*_0_(*t*) and the lethality rate of 2.0%, the simulation results of the model for 110 days are shown in Figure 4. The number of tested cases stabilized around 377000, while deaths accumulated around 35000. Again, we should note that the basic reproduction rate (*R*_0_(*t*)) varies with social behavior attitudes, such as social isolation policies and use of homemade masks, which may increase if people do not obey the right practices or decrease if the policies are intensified and respected by the population. Although it is a projection, considering the current trend that can change according to social behavior, the projected cases serve as estimated information for logistical decisions. Between these, provisioning beds for hospitalization of severe and critical cases (6.1%), the number of kits for tests, as well as morgue infrastructure, taking into account that the mortality rate, currently estimated at 2%, may increase as the adequate treatment capacity is saturated and the public health collapses.

**Figure 4:**
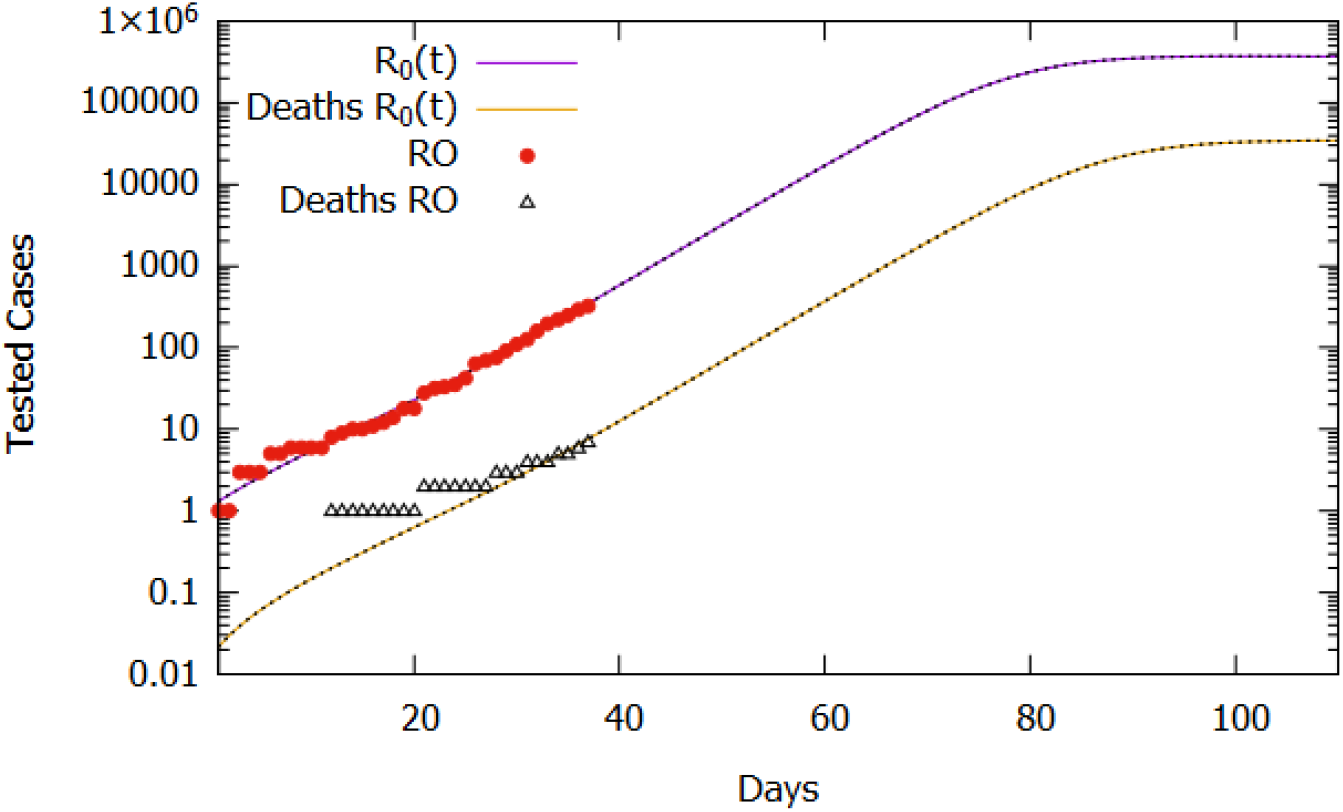
*Comparison between confirmed cases and the simulation for R_0_(t) according to eq. (2) for 110 days*.

As shown in Figure 1, for the model used, the increase in the basic reproduction rate causes an increase in the accumulated number of tested cases and, consequently, deaths, maintaining the same criteria and proportion of testing and estimated mortality rate. To assess the effect of the influence of the basic reproduction rate on the calibrated model, three cases and values of *R*_0_ (*t*) for *t ≥* 31 were established: maintenance of current isolation attitudes (*R*_0_(*t*) = 2.6), relaxation (*R*_0_(*t*) = 2.8) and intensification (*R*_0_(*t*) = 2.0). Table 1 shows that small changes in the basic reproduction rate, after the 31st day, result in significant changes in the number of days required to reach the peak of active cases, demonstrating the need for social isolation and use of homemade masks to reduce and delay the peak of new cases, as shown in Figure 5, in the absence of new treatments or vaccines against the virus.

**Table 1:** *Days for the occurrence of active cases peak as a function of the Basic Reproduction Rate from the 31st day*.

| $R_0(t)$ | Days | Peak |
| --- | --- | --- |
| 1,4 | 345 | 2741 |
| 1,5 | 238 | 5621 |
| 1,7 | 159 | 11630 |
| 2.0 | 115 | 19725 |
| 2,6 | 83 | 32764 |
| 2,8 | 78 | 36976 |

**Figure 5:**
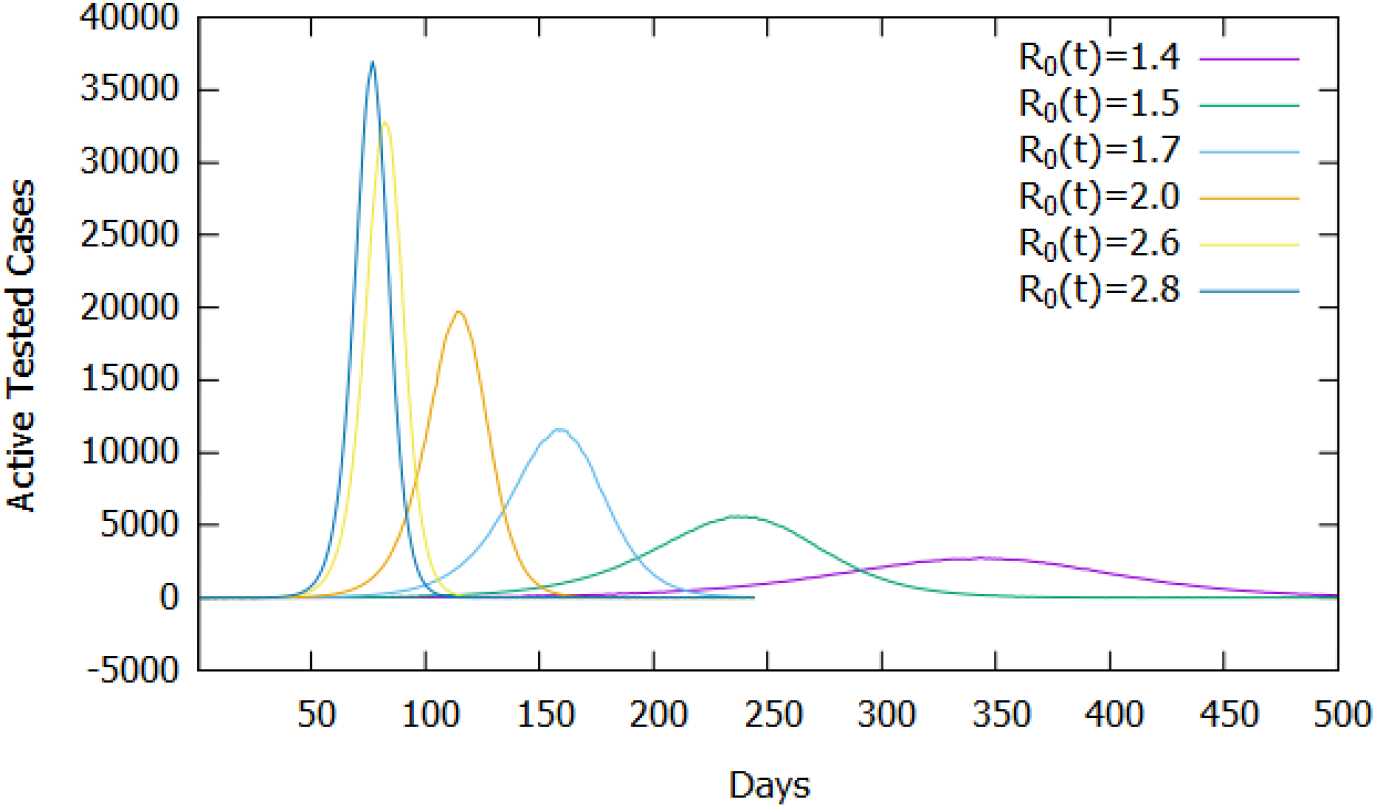
*Number of active cases according to the Basic Reproduction Rate (R_0_(t)) as of the 31st day*.

Another critical parameter is the proportion of testing (*ρ*) because, in the model, it represents the proportion of infected people tested and submitted to quarantine, preventing their contact with the rest of the population. Here, we estimated it at 20% due to the criteria and prevalence of severe cases and critics of COVID-19 reported in the literature. We have assessed this testing parameter influence and, consequently, the influence of the testing criteria in the accumulated cases obtained by the model; the parameter varied between 0.1 and 0.65. Figure 6 shows the results, where we observed that as the proportion of infected people tested increases to *ρ* = 0.4, there is a slight increase in the number of accumulated cases, this is due to a direct increase in confirmations without a significant impact on the effect of isolation caused by confirmations. For *ρ >* 0.4, the number of accumulated cases decreases, since, in mathematical modeling, after confirmation of infection, the individual is segregated from the population preventing the transmission of COVID-19. This decreasing rate of effective contact evidences, in the model, the need to change the testing criteria seeking to increase the proportion of known infected individuals and isolating them. One should note that for the proposed model, as shown in Figure 6, the most considerable decline in the number of accumulated cases starts at approximately *ρ* = 0.56, providing a practical scientific parameter for the establishment of new testing strategies and criteria established together with logistical restrictions of availability and costs. Despite testing costs, from an economic point of view, the increase in the proportion of testing can be an essential tool in monitoring and implementing selective isolation.

**Figure 6:**
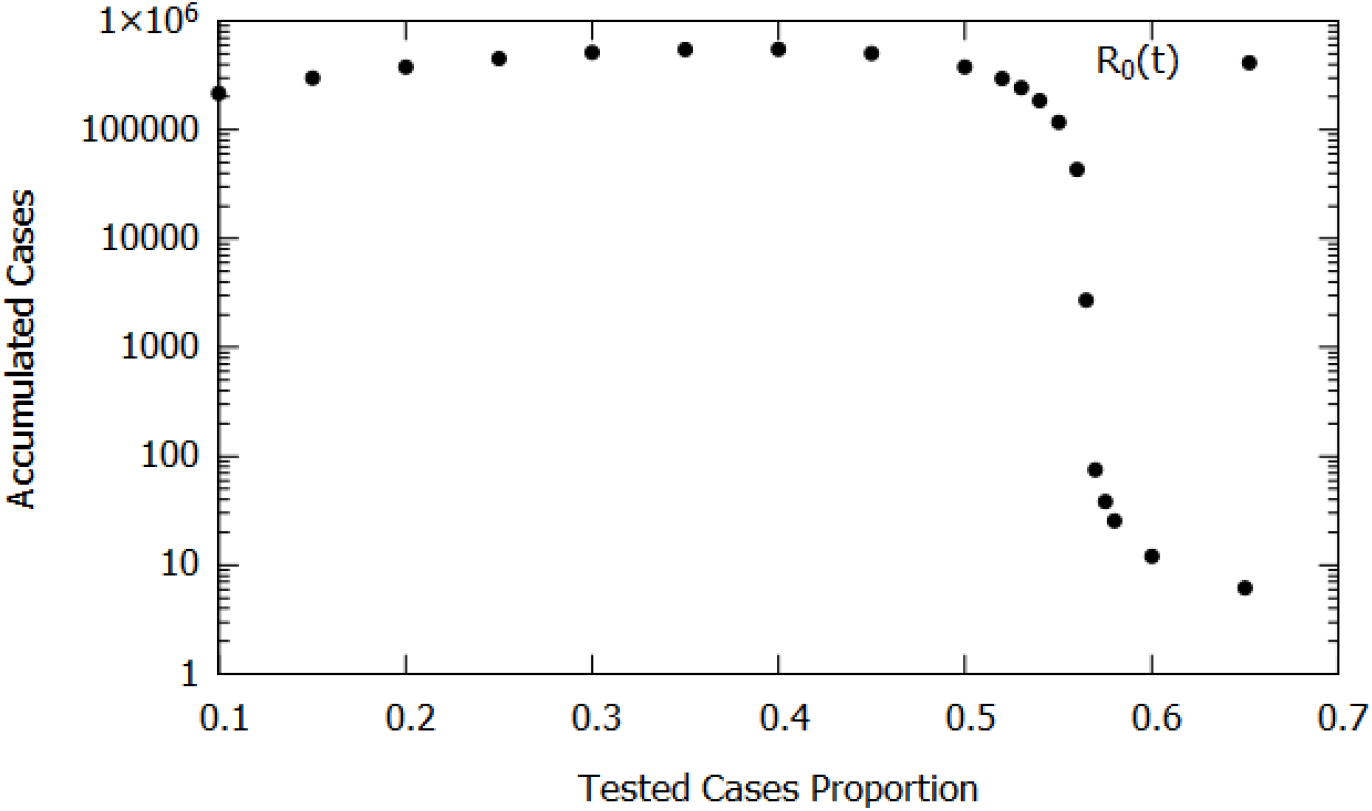
*Numbers of tested cases according to the proportion of tested cases*.

## 5. Final Remarks

The mathematical model proposed in this work to describe the evolution of COVID-19 in the State of Rondônia, Brazil, having considered the average periods of infection, incubation, and testing criteria obtained in the international literature, demonstrated good qualitative and quantitative adherence to the official data of confirmed cases.

In this model, the basic reproduction rate (*R*_0_) varies with time due to the social behavior of individuals, decreasing with the increase in isolation or the use of homemade masks, and being influenced by the logistical flow of testing, as logistical availability of test kits. Still, the temporal evolution of the number of tested and untested individuals are counted separately, according to the testing criteria established to consider the effect of the testing proportion.

The results show that the basic reproduction rate did not remain constant during the analyzed period, showing variations in social behavior and variations in the flow of testing or in the flow of official daily records, something we cannot estimate with the official information available. The temporal projections showed that the decrease in the basic reproduction rate for the next days, after the recorded data period, has a significant effect on the peak of active cases, as well as in the period in which it will occur in the future.

Regarding the proportion of testing, the results demonstrate the effectiveness of its increase beyond 40%, which causes a decrease in infected cases, since they can be immediately isolated, reducing the transmission speed. Thus, changing the testing criteria to include cases considered mild or moderate, easily confused, and described as asymptomatic, has beneficial effects. And, despite testing costs, increased testing can bring economic benefits if used in conjunction with selective isolation monitoring and implementation tools. During the period evaluated, according to the testing criteria, we conservatively estimated that the proportion of infected people tested was 20%, and according to the results obtained for the proposed model, its beneficial effects become very significant from about 56% of the infected population, thus including, in addition to all severe and critical cases, more mild and moderate cases in the testing criteria.

In conclusion, despite the limitations and simplifications inherent to any process of mathematical modeling of natural phenomena, the proposed model can be used as a basis to assist the planning of intensification or relaxation actions of social isolation to match the demand for treatment with infrastructure and adequate service capacity.

## Data Availability

All data are publicly available at https://covid.saude.gov.br/

